# Bayesian spatial modelling of childhood cancer incidence in Switzerland using exact point data: A nationwide study during 1985-2015

**DOI:** 10.1101/19001545

**Authors:** Garyfallos Konstantinoudis, Dominic Schuhmacher, Roland A Ammann, Tamara Diesch, Claudia E Kuehni, Ben D Spycher, for the Swiss Paediatric Oncology Group and the Swiss National Cohort Study Group

**Affiliations:** Institute of Social and Preventive Medicine (ISPM), University of Bern, Bern, Switzerland; Institute for Mathematical Stochastics, University of Göttingen, Germany; Department of Paediatrics Inselspital, Bern University Hospital, University of Bern, Bern, Switzerland; University Children’s Hospital Basel, Division of Pediatric Oncology/Hematology, Basel, Switzerland

**Keywords:** Cancer clusters, Medulloblastoma, Conditional autoregressive models, Gauss Markov random fields, point processes

## Abstract

**Background:** The aetiology of most childhood cancers is largely unknown. Spatially varying environmental factors such as traffic-related air pollution, background radiation and agricultural pesticides might contribute to the development of childhood cancer. We investigated the spatial variation of childhood cancers in Switzerland using exact geocodes of place of residence.

**Methods:** We included 5,947 children diagnosed with cancer during 1985-2015 at age 0-15 from the Swiss Childhood Cancer Registry. We modelled cancer risk using log-Gaussian Cox processes and indirect standardization to adjust for age and year of diagnosis. We examined whether the modelled spatial variation of risk can be explained by ambient air concentration of NO_2_, natural background radiation, area-based socio-economic position (SEP), linguistic region, years of existing general cancer registration in the canton or degree of urbanization.

**Results:** For all childhood cancers combined, the posterior median relative risk (RR), compared to the national level, varied by location from 0.83 to 1.13 (min to max). Corresponding ranges were 0.96 to 1.09 for leukaemia, 0.90 to 1.13 for lymphoma, and 0.82 to 1.23 for CNS tumours. The covariates considered explained 72% of the observed spatial variation for all cancers, 81% for leukaemia, 82% for lymphoma and 64% for CNS tumours. There was evidence of an association of background radiation and SEP with incidence of CNS tumours, (1.19;0.98-1.40) and (1.6;1-1.13) respectively.

**Conclusion:** Of the investigated diagnostic groups, childhood CNS tumours show the largest spatial variation in Switzerland. The selected covariates only partially explained the observed variation of CNS tumours suggesting that other environmental factors also play a role.

## Introduction

The causes of childhood cancers are poorly understood. Epidemiological research on the atomic bomb survivors indicated that ionising radiation in high doses can cause childhood leukaemia and central nervous system (CNS) tumours [1, 2]. A number of environmental factors have been suggested that could partially explain cancer risks in the general population, including traffic-related air pollution [3], background radiation [2, 4] and agricultural pesticides [5]. These risk factors vary in space and it is thus natural to expect spatial variation in childhood cancer incidence. Conversely, investigating the spatial variation of childhood cancer incidence might help generate new hypotheses about environmental risk and identify areas of potential environmental contamination.

Disease mapping, i.e. smoothing and visualising disease risk in space, is a common way of capturing the spatial variation of a disease. Several previous studies have investigated spatial variation in childhood cancer risk using disease mapping. Studies have focused on childhood leukaemia reported evidence of spatial variation in Ohio, Texas, Yorkshire [6-8], but not in France [9]. The study in Texas also examined childhood lymphomas and reported some evidence of spatial variation of Hodgkin lymphoma [7]. A study in Kenya reported evidence of spatial variation of Burkitt’s lymphoma with higher rates in the northern part of the country [10]. A study in Florida focusing on childhood brain tumours reported some evidence of high excess risk in several non-adjacent counties [11].

The mixed results might reflect differences between the countries or methodological limitations. Most previous studies relied on areal data (data aggregated on administrative units) [7-9, 11-13]. Results from such studies depend on spatial unit selected, which is referred to as the Modifiable Areal Unit Problem [14]. Furthermore, associations between cancer incidence and environmental factors assessed at group level may be subject ecological fallacy, i.e. they may not correctly reflect underlying associations at the individual level [15]. The use of precise geocodes can overcome the aforementioned issues. In a simulation study, we showed that spatial modelling based on exact geocodes is more sensitive in identifying areas of higher risk compared to traditional disease mapping based on count data aggregated to small administrative areas [16]. To the best of our knowledge only one study in Ohio had available precise geocodes, but the authors did not attempt to explain the observed variation of childhood leukaemia risk by incorporating environmental exposures in the model [6]. Lastly, all the previous studies used geographical information about the place of diagnosis only. Children may be more susceptible to certain environmental exposures early in life and thus location of residence at birth may be more relevant [17].

In this nationwide study, we investigated the spatial variation of childhood cancers using precise locations of residence. We performed analysis using place of birth and diagnosis. We focused on the following main diagnostic groups: all childhood cancers, childhood leukaemia, lymphoma and CNS tumours and assessed the extent to which selected covariates could explain the observed spatial variation.

## Methods

### Study population

We retrieved children diagnosed with cancer in Switzerland during 1985-2015 at age 0-15 from the Swiss Childhood Cancer Registry (SCCR). SCCR is a nationwide registry with high completeness. Estimates suggest that it includes 91% of all incident cases for the period 1985-2009 and >95% for 1995-2009.[18] It collects residential addresses from time of diagnosis back to birth. The addresses were geocoded according to the Swiss grid coordinate system using a combination of different sources of georeferenced building addresses including the Swiss postal system, the geoportal maintained by the Federal Office of Topography and Google Maps.

Population data was available through the Swiss National Cohort (SNC) which includes geocoded residential locations of all Swiss residents at time of censuses (1990, 2000 and 2010-2015). To calculate population at risk by age group, year and spatial unit (1km grid cell or municipality), we performed linear interpolation of age, year and spatial unit specific weights, see Additional File Text S1 and Figures S1-2. We then performed indirect standardization by calculating the expected number of cases adjusted by age and year: Let *q*_*i,j*_ be the nationwide cancer incidence and *P*_*i,j,k*_ the population counts with subscript referring to the *i*-th age group (0-4, 5-9, 10-15), *j*-th year (1985-2015), and *k*-th spatial unit (grid cell, or municipality). Then the expected number of cases in the *k*-th spatial unit is:

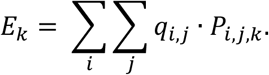

To calculate the expected number of cases for the analysis based on the location at birth we used a similar approach (Additional File Text S1).

### Outcomes

The SCCR classifies diagnoses according to the International Classification of Childhood Cancers Third Edition (ICCC3). We examined all childhood cancers combined (ICCC3 main groups I-XII) and then separately childhood leukaemia (ICCC3 main group I), lymphoma (ICCC3 main group II) and CNS tumours (ICCC3 main group III). We focused on the main diagnostic groups because of the larger sample size.

### Covariates

As potential explanatory variables, we included predicted ambient air concentration of NO_2_, predicted total dose rate from terrestrial gamma and cosmic radiation, neighbourhood-level socio-economic position (Swiss-SEP) [19], years of general cancer registration in the canton, language region and the degree of urbanisation as covariates (Table S1 and Figures S3-8 on the Additional File). Traffic-related air pollution and total background radiation were previously found to be associated with childhood cancer risks in Switzerland [20, 21]. We included SEP, linguistic region and degree of urbanisation to account for regional, socio-economic and socio-cultural differences. We included years of cantonal cancer registration to account for heterogeneous registry completeness. The SCCR records childhood cancer cases treated in one of the nine specialised paediatric oncology (SPOG) clinics and complements the registry with any additional cases recorded by the cantonal registries. Some cantons already had a cancer registry at the beginning of our study period, others established one during the study period and others after the end of the study. For cantons with more years of general registration, we thus expect the “apparent” childhood cancer incidence over the study period to be slightly higher.

### Statistical Analysis

We used log-Gaussian Cox processes (LGCPs) to model locations of incident cancer cases. A detailed description is provided in the Supplementary Text S2 [22]. Conditional on the risk surface, the point process assumed to generate the case locations is an inhomogeneous Poisson process. We model the continuous log-risk surface via a spatial mixed effects model, adjusting for covariate effects. The spatial variation is modelled as a random process *Z*(*s*), which is assumed to be a realization of a zero mean Gaussian random field with a Matérn covariance function and smoothness parameter *v* fixed to 1. The Gaussian field is then defined by two parameters, a variance parameter *σ*^2^ and a range parameter *ρ* (a distance at which the correlation between two points of the field is approximately 0.10). We fitted the model using the stochastic partial differential equation approach to approximate the continuous Gaussian field [23], and the Integrated Nested Laplace Approximation to perform accurate and computationally feasible Bayesian analysis [24, 25].

We computed maps of posterior median (unadjusted or adjusted for the covariates) of spatial relative risk (RR, i.e. exp{*Z*(*s*)}) compared to national level on a 1×1*km*^2^ grid. We also mapped exceedance probabilities defined as the posterior probability, in each grid cell, that RR exceeds 1. The fixed effects *β*_*i*_ (log-relative-risk per unit increase in the covariate) are reported as posterior median of RR, i.e. exp{*β*_*i*_}, and 95% credibility intervals (CI). The continuous variables NO_2_, ionizing radiation, SEP and years of cantonal cancer registration were scaled and thus exp{*β*_*i*_} is interpreted as the multiplicative change of the risk if at a fixed location when the covariate is increased by 1 standard deviation (SD).

They were included as linear terms since there was no indication for a more complex model (Additional File Figure S9). Henceforth, the model adjusted for the aforementioned covariates is referred to as the adjusted model, whereas the model without covariates as the unadjusted. Both adjusted and unadjusted models are standardized for population, age and year of diagnosis by including the expected number of cases as an offset in the model (Additional File Text S1-2).

We also report the percentage of variance explained by the selected risk factors by evaluating median and 95% CI of the posterior of an extension of Bayesian *R*^2^ [26]:

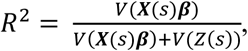

where *V*(·) denotes the variance over the *K* spatial units, ***β*** is the vector of intercept and covariates and ***X***(*s*) is the design matrix. We calculated *R*^2^ for the fully adjusted model, a model including all selected covariates except years of cantonal cancer registration (we refer to this set of covariates as ‘putative risk factors’), and the univariable model including only years of cantonal cancer registration. This allows us to distinguish spatial variation explained purely by the degree of completeness of registration from variation explained by covariates that might reflect aetiological factors (putative risk factors). For consistency with the literature, we also fitted the Besag-York-Mollié (BYM) model using disease counts per municipality, for more information see [27-29] and Text S2 of the Additional File.

### Sensitivity Analysis

We ran a sensitivity analysis to examine the robustness of the results with respect to different scalings of the penalized complexity priors for the range parameter of the latent field [28], with median range fixed at 1,10,60,120 and 240km.

## Results

### Study Population

We identified 5,969 cases with childhood cancer during 1985-2015 in Switzerland. We excluded 22 (0.3%) cases without available geocode of residence at diagnosis. Of the included 5,947 children, 32% (N = 1,880) had leukemia, 13% (N = 772) lymphoma and 22% (N = 1,290) a CNS tumor. For the analysis using location at birth we first excluded 1,194 cases born before 1985 and then 577 additional cases with no geocode at birth yielding 4,198 cases for the analysis (Table 1). Of the excluded cases for this analysis, 342 were born abroad, 114 were born in Switzerland but no address was recorded, while for 121 the country of birth was missing. The age and sex distribution follows similar patterns as in neighboring countries (Table 1) [30, 31].

**Table 1.**
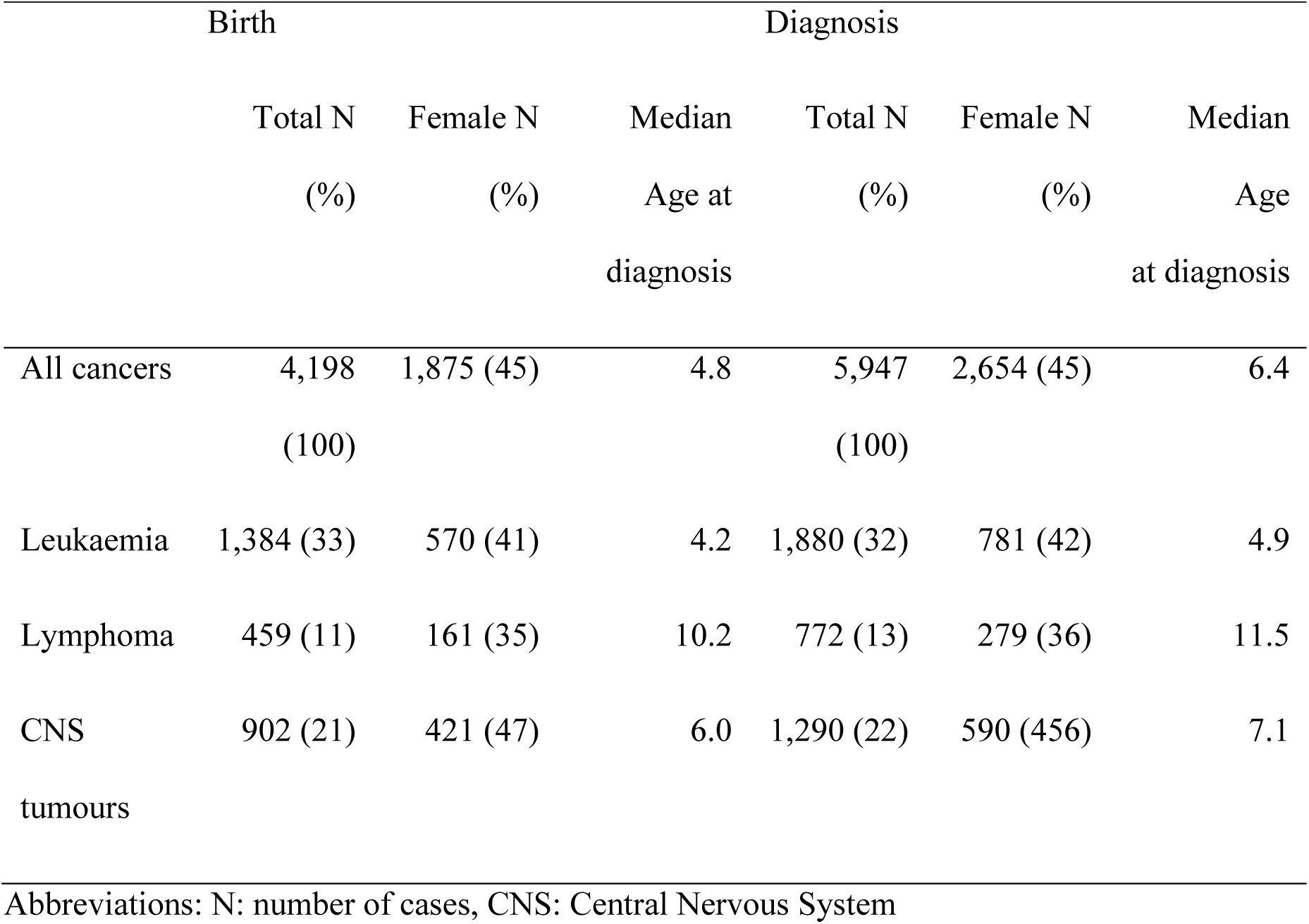
Number of cases and median age at diagnosis for the analysis based on the location at birth and diagnosis.

### Spatial analysis

We found evidence of spatial variation for all cancers combined and CNS tumours at diagnosis, Figure 1, Table 2 and Table S2. For leukaemia and lymphoma the posterior median of the variance hyperparameter of the Gaussian field (*σ*^2^) was shrunk to 0 or values close to 0, indicating small, if any, spatial variation (Table 2 and S2).

**Fig 1.**
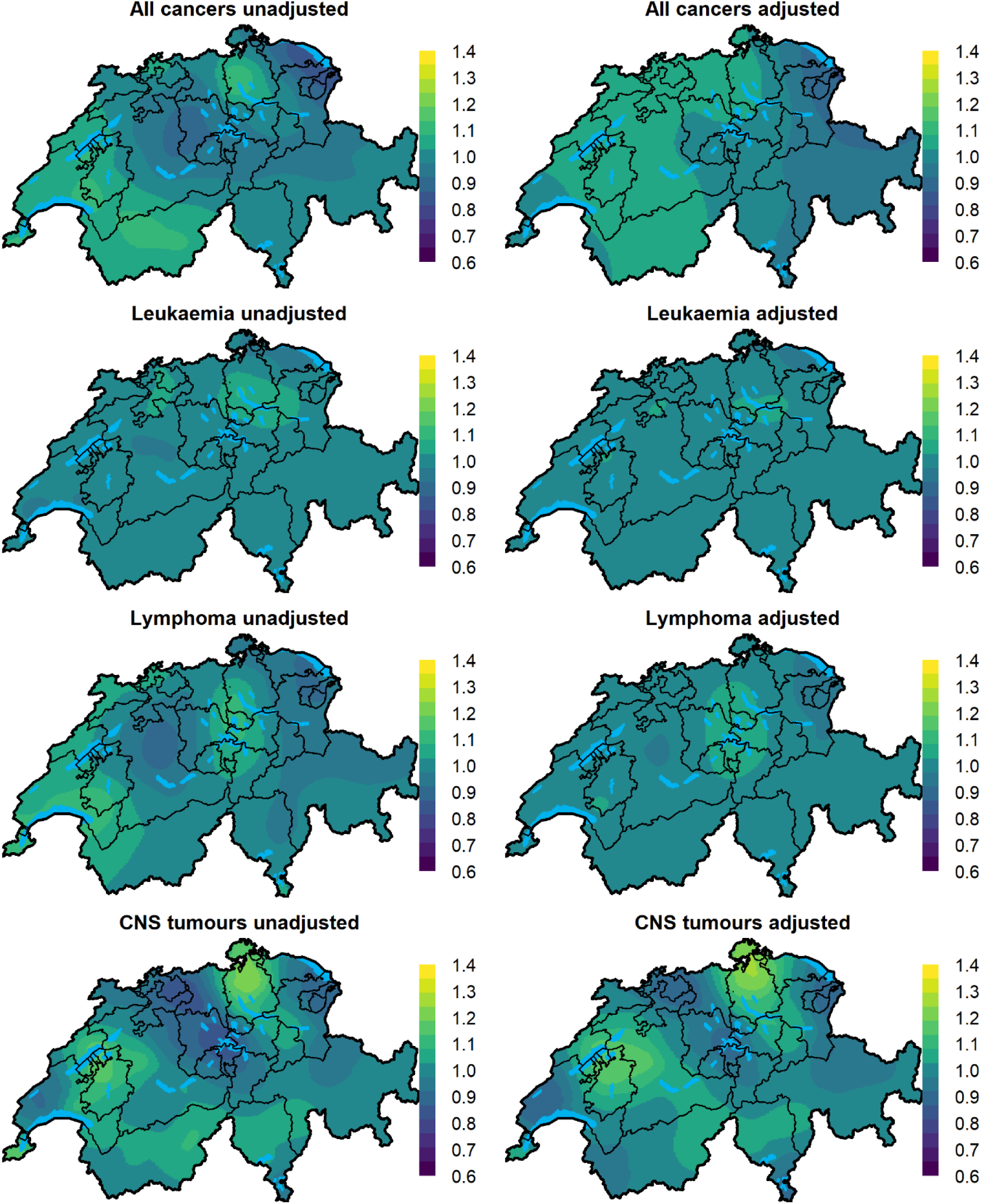
Maps of the median prosterior of the spatial relative risk for different cancer types during 1985-2015 in Switzerland. The adjusted models are models adjusted for NO_2_ total background radiation, SEP, years of cantonal registry, language region and level of urbanization.

**Table 2.**
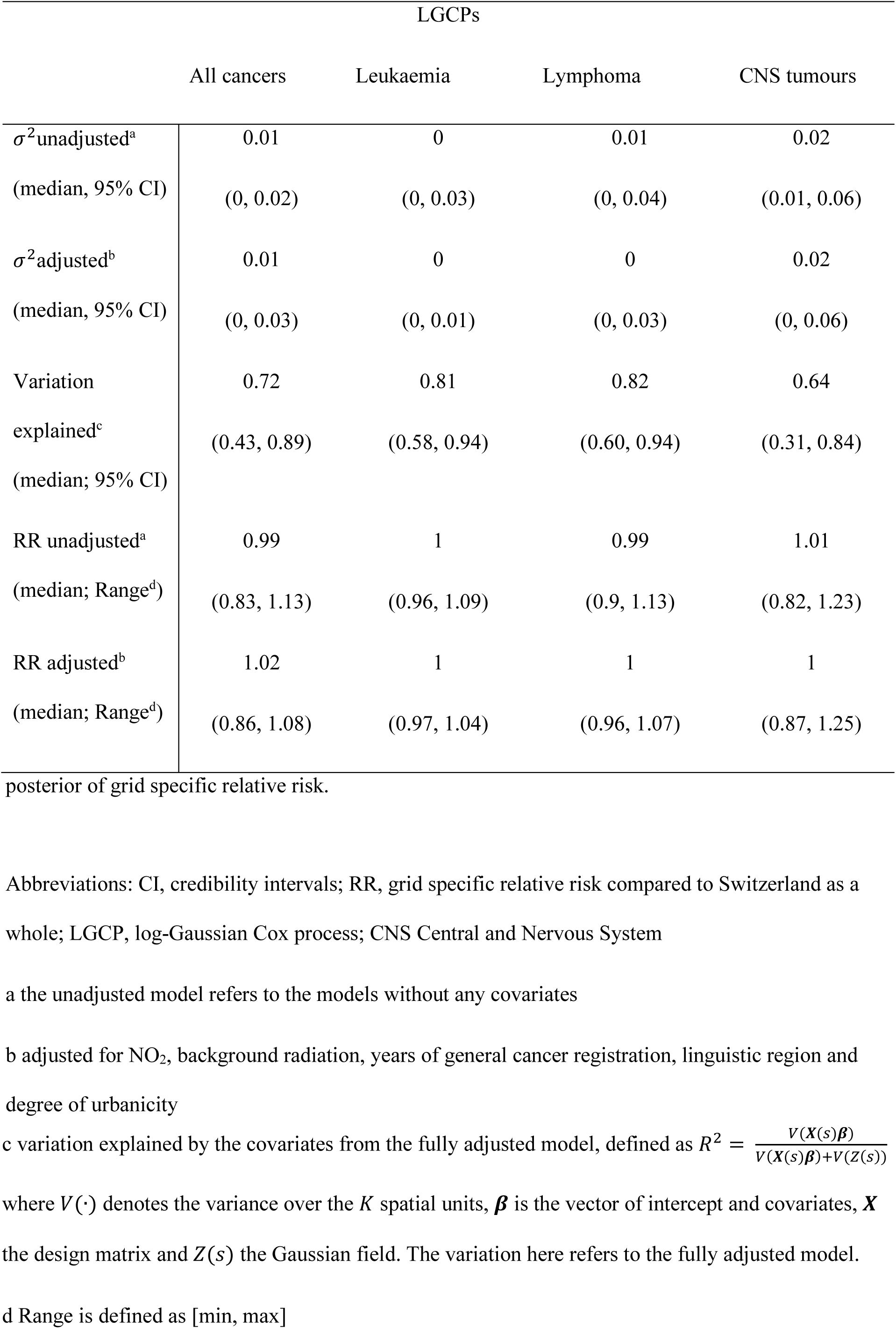
Median posterior of the variance hyperparameter of the Gaussian field (*σ*^2^) for the unadjusted and adjusted model, median posterior of variation explained (*V*(*Z*(*s*))) and median

|  | LGCPs |  |  |  |
| --- | --- | --- | --- | --- |
|  | All cancers | Leukaemia | Lymphoma | CNS tumours |
| $\sigma^2$unadjusted<sup>a</sup> | 0.01 | 0 | 0.01 | 0.02 |
| (median, 95% CI) | (0, 0.02) | (0, 0.03) | (0, 0.04) | (0.01, 0.06) |
| $\sigma^2$adjusted<sup>b</sup> | 0.01 | 0 | 0 | 0.02 |
| (median, 95% CI) | (0, 0.03) | (0, 0.01) | (0, 0.03) | (0, 0.06) |
| Variation explained<sup>c</sup> | 0.72 | 0.81 | 0.82 | 0.64 |
| (median; 95% CI) | (0.43, 0.89) | (0.58, 0.94) | (0.60, 0.94) | (0.31, 0.84) |
| RR unadjusted<sup>a</sup> | 0.99 | 1 | 0.99 | 1.01 |
| (median; Range<sup>d</sup>) | (0.83, 1.13) | (0.96, 1.09) | (0.9, 1.13) | (0.82, 1.23) |
| RR adjusted<sup>b</sup> | 1.02 | 1 | 1 | 1 |
| (median; Range<sup>d</sup>) | (0.86, 1.08) | (0.97, 1.04) | (0.96, 1.07) | (0.87, 1.25) |

posterior of grid specific relative risk.
Abbreviations: CI, credibility intervals; RR, grid specific relative risk compared to Switzerland as a whole; LGCP, log-Gaussian Cox process; CNS Central and Nervous System
a the unadjusted model refers to the models without any covariates
b adjusted for NO<sub>2</sub>, background radiation, years of general cancer registration, linguistic region and degree of urbanicity
- 1 c variation explained by the covariates from the fully adjusted model, defined as $R^2 = \frac{V(X(s)\boldsymbol{\beta})}{V(X(s)\boldsymbol{\beta}) + V(Z(s))}$ - 2 where $V(\cdot)$ denotes the variance over the $K$ spatial units, $\boldsymbol{\beta}$ is the vector of intercept and covariates, $\mathbf{X}$ - 3 the design matrix and $Z(s)$ the Gaussian field. The variation here refers to the fully adjusted model. - 4 d Range is defined as [min, max] - 5

For all cancers grouped together, the medians of the posterior distributions of RR evaluated at the centroids of 1×1*km*^2^ grid cells varied from 0.83 to 1.13 (min to max) throughout Switzerland, indicating at most a 13% increase in the risk in certain grid cells compared to Switzerland as a whole (Table 2 and Figure 1). The corresponding exceedance probability maps show areas, for which the posterior probability of having an RR greater than 1 is above 0.80 highlighted in light green or yellow (Figure 2). When we adjust for the selected covariates almost 72% (95% CI: 43%, 89%) of the observed variation was explained, with the median residual RR after adjustment varying from 0.86 to 1.08 (min to max), Figure 1, Table 2. The putative risk factors explained 65% (35%, 86%) of the observed variation (Additional File, Table S3). In the fully adjusted model, the factors NO_2_ (RR 1.02; 95% CI 0.99-1.06 per 1 SD increase in NO_2_), total background radiation (1.08; 0.99-1.18) and years of cantonal cancer registry (1.06; 1.03-1.09) were positively associated with cancer risk, whereas the association with the other covariates was weak (Figure 3 and Additional File Table S4).

**Fig 2.**
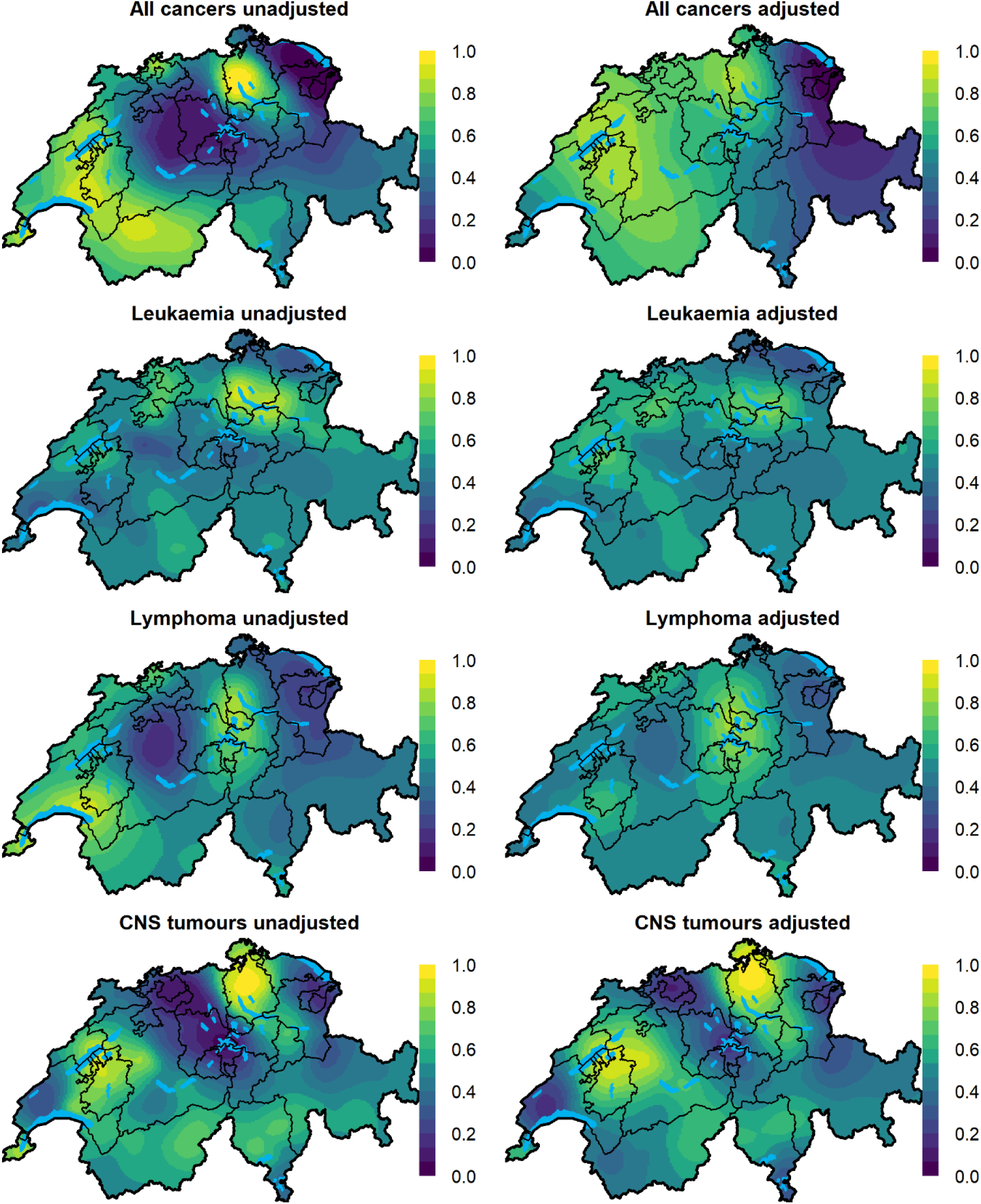
Maps of posterior probabilities that the spatial relative risk risk per grid cell is larger than 1 (exceedance probabilities) for different chilhdood cancers groups during 1985-2015 in Switzerland. The adjusted models are adjusted for NO_2_, total background radiation, SEP, years of cantonal registry, language region and level of urbanization.

**Fig 3.**
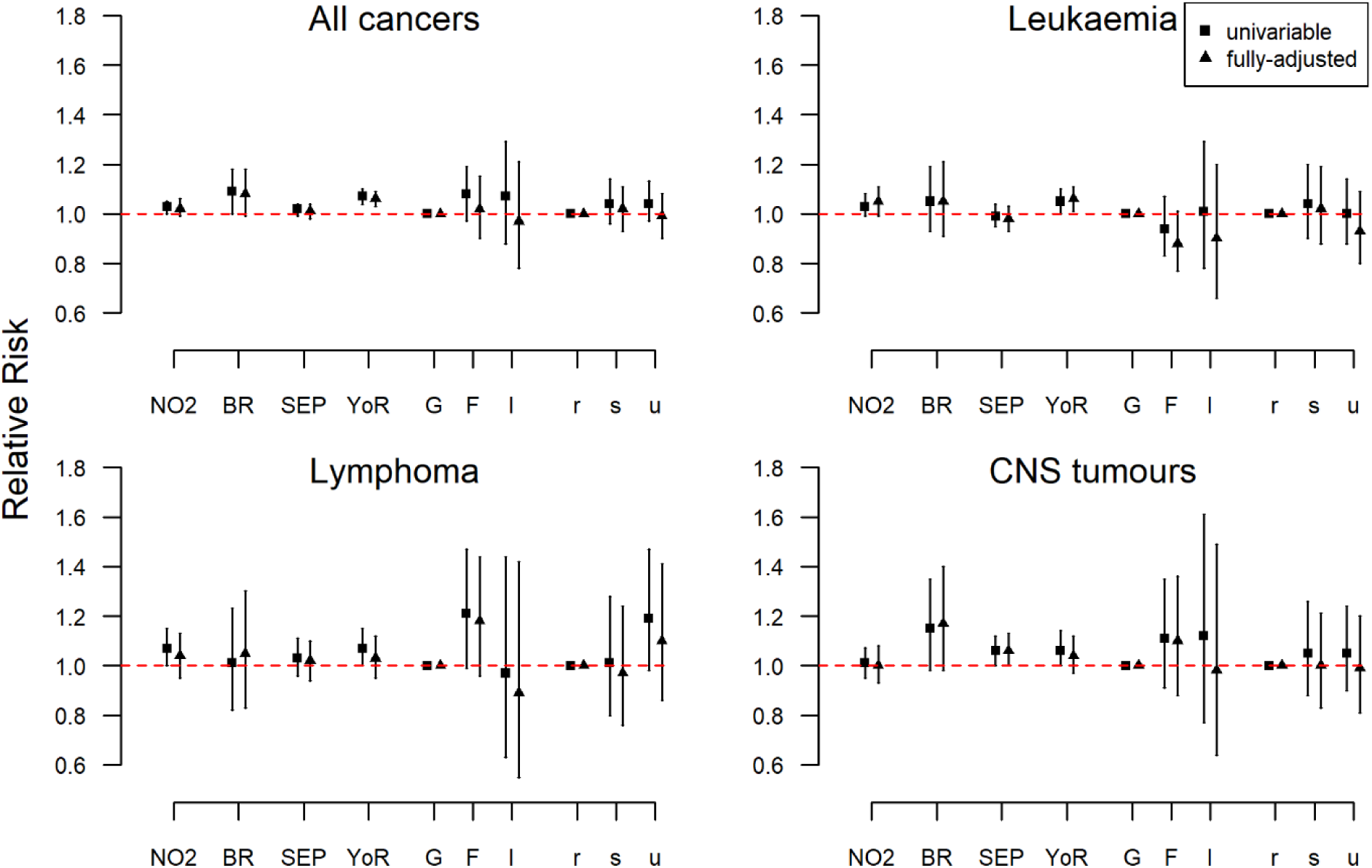
Univariable and fully adjusted regression analysis at time of diagnosis. The fixed effects are summarized using the posterior median of the relative risk together with 95% credibility regions. Abbrevations: NO_2_: Nitrogen Dioxide, CNS: Central Nervous System tumours, BR: Total dose background radiation, SEP: Socio-Economic Position, YoR: years of existing cantonal registry, G: German speaking part, F: French speaking part, I: Italian speaking part, r: rural areas, s: semi-urban areas, u: urban areas NO_2_, total background radiation, SEP and years of cantonal registry were scaled so that the standard deviations (SD) are 1 and considered as linear effects. Their interpretation is a multiplicative increase (or decrease) in the number of observed cases compared to the number of the expected cases per 1 SD increase (or decrease) in the covariate. The sd for NO_2_ is 77.7 *μg*/*m*^3^×10, for total background radiation 60.2 *nSv*/*h*, for SEP 8.7 units and for years of cantonal registry 11.6 years. The fully-adjusted models are models adjusted for NO_2_, total background radiation, SEP, years of cantonal registry, language region and level of urbanization.

Childhood leukaemia risks showed smaller spatial variation with the median posterior RR per grid cell varying from 0.96 to 1.09 on the unadjusted and from 0.97 to 1.04 on the fully adjusted model, Figure 1 and Table 2. The proportion of spatial variation explained by the selected covariates was 81% (58%, 94%), Table 2, whereas solely by the selected risk factor 64% (33%, 84%), Additional File, Table S3. In the fully adjusted model, the factors associated with the spatial risk of childhood leukaemia were NO_2_ exposure (1.05; 0.99-1.11) and years of cantonal cancer registry (1.06;1.01-1.11), Figure 3 and Additional File Table S4.

A small amount of spatial variation of RR was also observed for childhood lymphoma with the median RR varying from 0.90 to 1.13 on the unadjusted model and 0.96 to 1.07 on the adjusted model (Figure 1 and Table 2). About 82% (60%, 94%) of the observed spatial variation in the risk could be explained with the selected covariates, most of it due to the putative risk factors (Additional File, Table S3). In the fully adjusted model, the factor contributing most was living in the French speaking part of Switzerland with a 1.18 (0.96,1.44) RR increase compared to living in the German speaking part, Figure 3 and Additional File Table S4.

Among the investigated diagnostic groups, the greatest spatial variation of cancer risks was observed for childhood CNS tumours. The median posterior grid-specific RR varied from 0.82 to 1.23 before adjusting, and from 0.87 to 1.25 after adjusting for the selected covariates. These covariates explained 64% (31%, 84%) of the observed spatial variation, and the putative risk factors alone 62% (28%, 92%), Additional File, Table S3 and Table 2. Total background radiation exposure (1.17;0.98-1.4), SEP (1.06;1-1.13) and year of existing cantonal cancer registry (1.04;0.97-1.12) were positively associated with CNS tumour incidence. The association of the other covariates was weak, Figure 3 and Additional File Table S4.

We also examined the spatial variation of childhood cancers using place of birth. The spatial variation of cancer risks was generally smaller but the spatial patterns were largely consistent with the results for diagnosis (Additional File Figures S10-11 and Table S3 and S5).

We also examined the spatial variation using the BYM model. The maps and variation of median posterior RR were similar to the ones obtained by LGCPs, Additional File Figures S10-15. The estimates of the fixed effects were in the same direction but tended to be somewhat weaker than in the LGCP models (Additional File, Tables S4-7).

### Sensitivity analysis

The resulting maps and effect estimates varied only little when using different priors for the hyperparameters, Additional File Figures S16-25.

### Post-hoc analysis

Given the larger spatial variation in the risk of CNS tumours we ran several post-hoc analyses for this diagnostic group. First, we restricted the analysis to place of diagnosis for the period of 1995-2015 (n = 968), in which the coverage is highest (>95%). The resulting spatial pattern was closely similar to the main analysis (Figure S26). Second, we wanted to identify if the observed variation of CNS tumour was specific to particular diagnostic subgroups. We reran the analysis for place at diagnosis for astrocytoma (IIIb, n=511 cases), intracranial and intraspinal embryonal tumors (IIIc, n=266) and other CNS (IIIa, IIId-f, n=512), following the classification used in our previous analysis of spatial clustering of childhood cancers in Switzerland [16]. We found that intracranial and intraspinal embryonal tumors showed the highest spatial variation with the median posteriors RR varying from 0.74 to 1.59 (min to max) in the unadjusted and 0.74 to 1.38 in the adjusted model, Figure S27. However, the areas highlighted in Figures 1-2 stand out in all CNS subgroups Figures S27-28. Lastly, we hypothesized that differences in diagnostic practices between the nine SPOG clinics may explain the apparent spatial variation of CNS tumour risks. We thus constructed a spatial covariate reflecting the catchment areas of the different SPOG centres. Including an additional random effect to adjust for these catchment areas only slightly reduced the unexplained spatial variation. The spatial pattern of relative risk remained largely unchanged (Additional File Text S3 includes the analysis and figures).

## Discussion

### Main findings

This nationwide study based on precise locations of residence sheds new light on the spatial variation of childhood cancer incidence in Switzerland and the extent to which this variation can be explained by environmental exposures and other spatial covariates. The spatial variation of cancer risk was small for childhood leukaemia and lymphoma and mostly explained by covariates. That of CNS tumours, particularly intracranial and intraspinal embryonal tumours, was larger and persisted after adjustment for covariates. Duration of general cancer registration in the canton was associated with higher observed cancer risk. Other covariates associated with cancer incidence included ambient air concentration of NO_2_ for all cancers, lymphoma and leukaemia and SEP and dose rates from terrestrial gamma and cosmic background radiation for CNS tumours and all cancers.

### Comparison of our study with other spatial analyses of childhood cancer risks

Compared to other studies that have investigated the spatial distribution of childhood cancers, our study stands out in that it uses precise geocoded place of residence and attempts to explain any spatial variation with commonly discussed putative environmental risk factors and completeness of registration. Our study is comparable with studies that performed parametric disease mapping, and in the lack of other studies that used LGCPs, with the two previous studies that investigated the spatial variation of childhood leukaemia risks using areal data and BYM models [8, 9, 11]. A study in France on acute leukaemia reported no evidence of spatial variation in the incidence of acute leukaemia at the département level [9]. A study in Yorkshire using data aggregated on the electoral ward level reported higher childhood leukaemia risks in the less populated county of North Yorkshire [8]. We did not observe higher leukaemia risk in less populated areas. Our results are in agreement with a study in Florida that reported evidence of spatial variation of brain tumours for cases 0-19 years old [11].

Other studies examining the spatial distribution of childhood cancer have focused on extra-Poisson variation and spatial clustering [32]. The general picture shows mixed results for childhood leukaemia and weak or no evidence of spatial clustering of lymphoma and CNS tumours [33-35]. In previous studies using the same data, we found no evidence of clustering of childhood cancers, leukaemia, lymphoma or CNS tumours, but weak evidence, consistent with the literature, for Hodgkin lymphoma and embryonal CNS tumours [36, 37]. We observed a cluster of intracranial and intraspinal CNS tumours in the French speaking part of Switzerland consistent with the pattern observed for CNS tumours in the present study [36].

### Comparison of our study with other studies on environmental risk factors of childhood cancer

The observed spatial associations between childhood cancer risks and putative risk factors are in broad agreement with other studies that have investigated these associations disregarding the spatial context.

Of the included covariates in the current study, NO_2_ showed the strongest spatial association with childhood leukaemia risks. There is increasing evidence of a link between traffic related air pollution and childhood cancers, in particular childhood leukaemia [38]. In recent meta-analyses associations with leukaemia risks were strongest for exposure to benzene and weaker for NO_2_ [3]. Using partly overlapping data, we reported an increased risk of leukaemia among children living less than 100m from a highway [20].

Previous studies investigating childhood cancer risks in relation to background ionising radiation showed mixed results [21, 39-42]. While two studies reported associations between childhood leukaemia and gamma radiation [21, 39], others found no evidence of an association [40-42]. Using partly overlapping data, we previously reported evidence of associations with gamma radiation for both childhood leukaemia and CNS tumours [21]. In the current study the association was largest for all cancers and CNS tumours. The evidence from other studies examining the effect of gamma radiation on the risks of CNS tumours in children was weak [39, 41].

Our study found evidence of an association between SEP and CNS tumours. Previous studies in Switzerland have reported weak association between socioeconomic status and childhood leukaemia incidence, but a strong effect for CNS survival [43, 44]. Our results are consistent with a large UK case-control study which reported increased risk of CNS tumours in higher social classes [45]. A recent study in Spain also reported a positive association between risk of CNS tumours and socioeconomic status [46]. In contrast, a study in North-West England [47] and a study from Norway [48] found no evidence of an association between CNS tumours and measures of socio-economic status.

### Strengths and Limitations

To the best of our knowledge this is the first study attempting to model and explain the spatial distribution of childhood cancers using precise locations of residence. We used LGCPs, which represent the current state of the art for modelling such point data of disease incidence and, as we have recently shown, outperform traditional methods in identifying high risk areas [16]. These models allowed us to incorporate spatial covariates and quantify their contribution to explaining the observed spatial variation. We also tried to disentangle variation attributed to registration completeness from variation due to putative risk factors. Furthermore, in contrast to previous studies, we examined both place of birth and diagnosis. Although results were closely similar, this comparison could potentially have revealed differences in time windows of susceptibility to different risk factors. The population at risk was retrieved from national censuses and cases from a nationwide registry with high completeness [49]. We attempted to correct for potential selection bias due to regional differences in case ascertainment by including years of general cancer registration and, in post-hoc analysis, SPOG centre catchment areas.

Due to data availability, we could not include all potential environmental risk factors discussed in the literature, for instance pesticide exposure. Furthermore, the spatial covariates included are subject to measurement errors and do not perfectly capture the spatial variation of residential exposures. We had little information about the magnitude of measurement errors, making it hard to propagate it in our modelling framework. Although we partly adjusted for differences in registration coverage, there may still be differences unaccounted for by our analyses.

### Interpretation of findings

Although the overall completeness of SCCR is larger than 95% after the mid-90s [49], we found that years of existing cancer registry can influence the apparent spatial variation of childhood cancers based on data from SCCR. This suggests that there are regional differences in registration completeness, which should be accounted for in future aetiological studies in Switzerland.

Our results are suggestive of an environmental aetiology for childhood CNS tumours and of aetiological differences between their histological subtypes. In post-hoc analyses, the observed spatial variation was not fully explained by differences in cancer registration in the early years of the SCCR as it persisted in the more recent periods. Neither did differences between SPOG centres, for instance in ascertainment practices, explain the spatial variation. Unmeasured environmental risk factors are thus a likely explanation. Possibly, spatial differences in the prevalence of genetic syndromes associated with these tumours might also partially explain the observed variation. In future research, there should be increased attention on putative environmental risk factors of CNS tumours, including SEP, background radiation and pesticide exposure (which was not accounted for in our analyses).

## Conclusion

This study provides evidence of spatial differences in the incidence of childhood CNS tumours in Switzerland that could be partially explained by variations in socio-economic factors and natural background radiation. The spatial variation of the risks for childhood leukaemia and lymphoma was smaller and mostly explained by measured covariates. Our study provides further support for an environmental aetiology for childhood CNS tumours, highlighting the need for future studies to distinguish between histologic subtypes.

## Data Availability

The outcome data includes sensitive geographical information about children with cancer in Switzerland and thus cannot be made available.

## Abbreviations

CNS: Central nervous system
SCCR: Swiss childhood cancer registry
SNC: Swiss national cohort
ICCC3: International classification of childhood cancers 3^rd^ edition
NO_2_: Nitrogen dioxide
SEP: Socioeconomic position
SPOG: Swiss paediatric oncology group
LGCP: Log-Gaussian Cox process
RR: Relative risk
SD: Standard deviation
BYM: Besag-York-Mollié
UK: United Kingdom

## Acknowledgements

The work of the Swiss Childhood Cancer Registry is supported by the Swiss Paediatric Oncology Group (www.spog.ch), Schweizerische Konferenz der kantonalen Gesundheitsdirektorinnen und – direktoren (www.gdk-cds.ch), Swiss Cancer Research (www.krebsforschung.ch), Kinderkrebshilfe Schweiz (www.kinderkrebshilfe.ch), Ernst-Göhner Stiftung, Stiftung Domarena and National Institute of Cancer Epidemiology and Registration (www.nicer.org).

We thank the Swiss Federal Statistical Office for providing mortality and census data and for the support which made the Swiss National Cohort and this study possible. This work was supported by the Swiss National Science Foundation (grant nos. 3347CO-108806, 33CS30_134273 and 33CS30_148415).

Member of study groups

The members of the Swiss Pediatric Oncology Group Scientific Committee:

R A Ammann (Bern), K Scheinemann (Aarau), M Ansari (Geneva), M Beck Popovic(Lausanne), P Brazzola (Bellinzona), J Greiner (St. Gallen), M Grotzer (Zurich), H Hengartner (St Gallen), T Kuehne (Basel), J Rössler (Bern), F Niggli (Zurich), F Schilling (Lucerne), N von der Weid (Basel)

The members of the Swiss National Cohort Study Group:

Matthias Egger (Chairman of the Executive Board), Adrian Spoerri and Marcel Zwahlen (all Bern), Milo Puhan (Chairman of the Scientific Board), Matthias Bopp (both Zurich), Martin Röösli (Basel), Murielle Bochud (Lausanne) and Michel Oris (Geneva).

## Authors’ contribution

Conceptualisation: GK, BS; Methodology: GK, DS, BS; Formal analysis: GK; Validation: BS, DS, CK RA and TD; Writing—original draft: GK; Writing—review and editing: GK, DS, TD, RA, CK and BS; Resources: CK, TD, RA, BS; Supervision: DS and BS.

## Ethics approval and consent to participate

Ethics approval was granted through the Ethics Committee of the Canton of Bern to the SCCR on the 22th of July 2014 (KEK-BE: 166/2014).

## Consent of publication

Not applicable

## Data availability statement

The Swiss Childhood Cancer Registry is the permanent repository of data on childhood cancer cases used in this study. This data cannot be made publicly available for both legal and ethical reasons as this would compromise patient confidentiality and participant privacy. Interested researchers may contact the corresponding author or the Swiss Childhood Cancer Registry (http://childhoodcancerregistry.ch/) via its online contact form for further information.

## References

1. Wrensch M, Minn Y, Chew T, Bondy M, Berger MS: Epidemiology of primary brain tumors: current concepts and review of the literature. Neuro-oncology 2002, 4(4):278–299.

2. Wakeford R: The risk of childhood leukaemia following exposure to ionising radiation--a review. Journal of radiological protection : official journal of the Society for Radiological Protection 2013, 33(1):1–25.

3. Filippini T, Hatch EE, Rothman KJ, Heck JE, Park AS, Crippa A, Orsini N, Vinceti M: Association between Outdoor Air Pollution and Childhood Leukemia: A Systematic Review and Dose–Response Meta-Analysis. Environmental health perspectives 2019, 4(127):046002.

4. Little MP, Wakeford R, Borrego D, French B, Zablotska LB, Adams MJ, Allodji R, de Vathaire F, Lee C, Brenner AV et al: Leukaemia and myeloid malignancy among people exposed to low doses (< 100 mSv) of ionising radiation during childhood: a pooled analysis of nine historical cohort studies. Lancet Haematol 2018, 5(8):E346–E358.

5. Van Maele-Fabry G, Gamet-Payrastre L, Lison D: Residential exposure to pesticides as risk factor for childhood and young adult brain tumors: A systematic review and meta-analysis. Environment international 2017, 106:69–90.

6. Wheeler DC: A comparison of spatial clustering and cluster detection techniques for childhood leukemia incidence in Ohio, 1996-2003. International journal of health geographics 2007, 6:13.

7. Thompson JA, Carozza SE, Zhu L: An evaluation of spatial and multivariate covariance among childhood cancer histotypes in Texas (United States). Cancer Causes & Control 2007, 18(1):105–113.

8. Manda SO, Feltbower RG, Gilthorpe MS: Investigating spatio-temporal similarities in the epidemiology of childhood leukaemia and diabetes. European journal of epidemiology 2009, 24(12):743.

9. Faure Cabc, Mollie Ad, Bellec Sabc, Guyot-Goubin Aabc, Clavel Jabc, Hemon Dab: Geographical variations in the incidence of childhood acute leukaemia in France over the period 1990-2004. European Journal of Cancer Prevention 2009, 18(4):267–279.

10. Rainey JJ, Omenah D, Sumba PO, Moormann AM, Rochford R, Wilson ML: Spatial clustering of endemic Burkitt’s lymphoma in high-risk regions of Kenya. International Journal of Cancer 2007, 120(1):121–127.

11. Lawson A, Rotejanaprasert C: Childhood Brain Cancer in Florida: A Bayesian Clustering Approach (vol 1, pg 99, 2014). Statistics and Public Policy 2015, 2(1):93–93.

12. Ortega-García J, López-Hernández F, Cárceles-Álvarez A, Santiago-Rodríguez E, Sánchez A, Bermúdez-Cortes M, Fuster-Soler J: Analysis of small areas of paediatric cancer in the municipality of Murcia (Spain). Anales de Pediatría (English Edition) 2016, 84(3):154–162.

13. Torabi M, Rosychuk RJ: An examination of five spatial disease clustering methodologies for the identification of childhood cancer clusters in Alberta, Canada. Spatial and spatiotemporal epidemiology 2011, 2(4):321–330.

14. Openshaw S: The Modifiable Areal Unit Problem: Geo Books; 1984.

15. Wakefield J: Ecologic studies revisited. Annu Rev Public Health 2008, 29:75–90.

16. Konstantinoudis G, Schuhmacher D, Rue H, Spycher B: Discrete versus continuous domain models for disease mapping. ArXiv preprint 180804765v1 2018.

17. Anderson LM, Diwan BA, Fear NT, Roman E: Critical windows of exposure for children’s health: cancer in human epidemiological studies and neoplasms in experimental animal models. Environmental health perspectives 2000, 108 Suppl 3:573–594.

18. Adam M, Schikowski T, Carsin AE, Cai Y, Jacquemin B, Sanchez M, Vierkotter A, Marcon A, Keidel D, Sugiri D et al: Adult lung function and long-term air pollution exposure. ESCAPE: a multicentre cohort study and meta-analysis. Eur Respir J 2015, 45(1):38–50.

19. Panczak R, Galobardes B, Voorpostel M, Spoerri A, Zwahlen M, Egger M, Swiss National C, Swiss Household P: A Swiss neighbourhood index of socioeconomic position: development and association with mortality. Journal of epidemiology and community health 2012, 66(12):61129–1136.

20. Spycher BD, Feller M, Roosli M, Ammann RA, Diezi M, Egger M, Kuehni CE: Childhood cancer and residential exposure to highways: a nationwide cohort study. European journal of epidemiology 2015, 30(12):1263–1275.

21. Spycher BD, Lupatsch JE, Zwahlen M, Roosli M, Niggli F, Grotzer MA, Rischewski J, Egger M, Kuehni CE, Swiss Pediatric Oncology G et al: Background ionizing radiation and the risk of childhood cancer: a census-based nationwide cohort study. Environmental health perspectives 2015, 123(6):622–628.

22. Moller J, Syversveen AR, Waagepetersen RP: Log Gaussian Cox processes. Scandinavian Journal of Statistics 1998, 25(3):451–482.

23. Lindgren F, Rue H, Lindstrom J: An explicit link between Gaussian fields and Gaussian Markov random fields: the stochastic partial differential equation approach. J R Stat Soc B 2011, 73:423–498.

24. Rue H, Martino S, Chopin N: Approximate Bayesian inference for latent Gaussian models by using integrated nested Laplace approximations. Journal of the royal statistical society: Series b (statistical methodology) 2009, 71(2):319–392.

25. Rue H, Riebler A, Sørbye SH, Illian JB, Simpson DP, Lindgren FK: Bayesian computing with INLA: a review. Annual Review of Statistics and Its Application 2017, 4:395–421.

26. Gelman A, Goodrich B, Gabry J, Vehtari A: R-squared for Bayesian Regression Models. The American Statistician 2018:1–7.

27. Besag J, York J, Mollié A: A Bayesian image restoration with two applications in spatial statistics. Ann Inst Statist Math 1991, 43:1–59.

28. Simpson D, Rue H, Riebler A, Martins TG, Sørbye SH: Penalising model component complexity: A principled, practical approach to constructing priors. Statistical Science 2017, 32(1):1–28.

29. Riebler A, Sorbye SH, Simpson D, Rue H: An intuitive Bayesian spatial model for disease mapping that accounts for scaling. Statistical methods in medical research 2016, 25(4):1145–1165.

30. Kaatsch P SC: German Childhood Cancer Registry - Report 2015 (1980-2014). Institute of Medical Biostatistics, Epidemiology and Informatics (IMBEI) at the University Medical Center of the Johannes Gutenberg University Mainz, Germany; 2015.

31. Lacour B, Guyot-Goubin A, Guissou S, Bellec S, Desandes E, Clavel J: Incidence of childhood cancer in France: National Children Cancer Registries, 2000-2004. Eur J Cancer Prev 2010, 19(3):173–181.

32. Bellec S, Hemon D, Rudant J, Goubin A, Clavel J: Spatial and space-time clustering of childhood acute leukaemia in France from 1990 to 2000: a nationwide study. British journal of cancer 2006, 94(5):763–770.

33. Armstrong BG: Effect of measurement error on epidemiological studies of environmental and occupational exposures. Occupational and environmental medicine 1998, 55(10):651–656.

34. McNally RJ, Eden TO: An infectious aetiology for childhood acute leukaemia: a review of the evidence. British journal of haematology 2004, 127(3):243–263.

35. Goujon S, Kyrimi E, Faure L, Guissou S, Hemon D, Lacour B, Clavel J: Spatial and temporal variations of childhood cancers: Literature review and contribution of the French national registry. Cancer medicine 2018.

36. Konstantinoudis G, Kreis C, Ammann RA, Niggli F, Kuehni CE, Spycher BD: Spatial clustering of childhood cancers in Switzerland: a nationwide study. Cancer Causes & Control 2018.

37. Konstantinoudis G, Kreis C, Ammann RA, Niggli F, Kuehni CE, Spycher BD, Swiss Paediatric Oncology G, the Swiss National Cohort Study G: Spatial clustering of childhood leukaemia in Switzerland: A nationwide study. International journal of cancer 2017, 141(7):1324–1332.

38. Filippini T, Heck JE, Malagoli C, Giovane CD, Vinceti M: A review and meta-analysis of outdoor air pollution and risk of childhood leukemia. Journal of environmental science and health Part C, Environmental carcinogenesis & ecotoxicology reviews 2015, 33(1):36–66.

39. Kendall GM, Little MP, Wakeford R, Bunch KJ, Miles JC, Vincent TJ, Meara JR, Murphy MF: A record-based case-control study of natural background radiation and the incidence of childhood leukaemia and other cancers in Great Britain during 1980-2006. Leukemia 2013, 27(1):3–9.

40. Demoury C, Marquant F, Ielsch G, Goujon S, Debayle C, Faure L, Coste A, Laurent O, Guillevic J, Laurier D et al: Residential Exposure to Natural Background Radiation and Risk of Childhood Acute Leukemia in France, 1990 - 2009. Environmental health perspectives 2016.

41. Spix C, Grosche B, Bleher M, Kaatsch P, Scholz-Kreisel P, Blettner M: Background gamma radiation and childhood cancer in Germany: an ecological study. Radiation and environmental biophysics 2017, 56(2):127–138.

42. Nikkila A, Erme S, Arvela H, Holmgren O, Raitanen J, Lohi O, Auvinen A: Background radiation and childhood leukemia: A nationwide register-based case-control study. International journal of cancer 2016, 139(9):1975–1982.

43. Adam M, Kuehni CE, Spoerri A, Schmidlin K, Gumy-Pause F, Brazzola P, Probst-Hensch N, Zwahlen M: Socioeconomic Status and Childhood Leukemia Incidence in Switzerland. Frontiers in oncology 2015, 5:139.

44. Adam M, Rueegg CS, Schmidlin K, Spoerri A, Niggli F, Grotzer M, von der Weid NX, Egger M, Probst-Hensch N, Zwahlen M et al: Socioeconomic disparities in childhood cancer survival in switzerland. International journal of cancer 2016.

45. Keegan TJ, Bunch KJ, Vincent TJ, King JC, O’Neill KA, Kendall GM, Maccarthy A, Fear NT, Murphy MF: Case-control study of paternal occupation and social class with risk of childhood central nervous system tumours in Great Britain, 1962-2006. British journal of cancer 2013, 108(9):1907–1914.

46. Ramis R, Tamayo-Uria I, Gomez-Barroso D, Lopez-Abente G, Morales-Piga A, Pardo Romaguera E, Aragones N, Garcia-Perez J: Risk factors for central nervous system tumors in children: New findings from a case-control study. PLoS One 2017, 12(2):e0171881.

47. McNally RJ, Alston RD, Eden TO, Kelsey AM, Birch JM: Further clues concerning the aetiology of childhood central nervous system tumours. European journal of cancer 2004, 40(18):2766–2772.

48. Del Risco Kollerud R, Blaasaas KG, Claussen B: Poverty and the risk of leukemia and cancer in the central nervous system in children: A cohort study in a high-income country. Scand J Public Health 2015, 43(7):736–743.

49. Schindler M, Mitter V, Bergstraesser E, Gumy-Pause F, Michel G, Kuehni CE, Swiss Paediatric Oncology G: Death certificate notifications in the Swiss Childhood Cancer Registry: assessing completeness and registration procedures. Swiss Med Wkly 2015, 145:w14225.

